# Vaccination of COVID-19 Convalescent Plasma Donors Increases Binding and Neutralizing Antibodies Against SARS-CoV-2 Variants

**DOI:** 10.1101/2021.10.28.21265622

**Authors:** Clara Di Germanio, Graham Simmons, Chloe Thorbrogger, Rachel Martinelli, Mars Stone, Thomas Gniadek, Michael P Busch

**Author notes:** Correspondence: M. P. Busch, Vitalant Research Institute, 270 Masonic Ave, San Francisco, CA 94118. these authors contributed equally to the manuscript (Clara Di Germanio, Graham Simmons).

## Abstract

**Background:** COVID-19 convalescent plasma (CCP) was widely used as passive immunotherapy during the first waves of SARS-CoV-2 infection in the US. However, based on observational studies and randomized controlled trials, beneficial effects of CCP were limited, and its use was virtually discontinued early in 2021, in concurrence with increased vaccination rates and availability of monoclonal antibody (mAb) therapeutics. However, as new variants of the SARS-CoV-2 spread, interest in CCP derived from vaccine-boosted CCP donors is resurging. The effect of vaccination of previously infected CCP donors on antibodies against rapidly spreading variants of concern (VOC) is still under investigation.

**Study Design/Methods:** In this study, paired samples from 11 CCP donors collected before and after vaccination were tested to measure binding antibodies levels and neutralization activity against the ancestral and SARS-CoV-2 variants (Wuhan-Hu-1, B.1.1.7, B.1.351, P.1, D614G, B.1.617.2, B.1.427) on the Ortho Vitros Spike Total Ig and IgG assays, the MSD V-PLEX SARS-CoV-2 Panel 6 arrays for IgG binding and ACE2 inhibition, and variant-specific Spike Reporter Viral Particle Neutralization (RVPN) assays.

**Results/Findings:** Binding and neutralizing antibodies were significantly boosted by vaccination, with several logs higher neutralization for all the variants tested post-vaccination compared to the pre-vaccination samples, with no difference found among the individual variants.

**Discussion:** Vaccination of previously infected individuals boosts antibodies including neutralizing activity against all SARS-CoV-2 VOC, including the current spreading delta (B.1.617.2) variant. Animal model and human studies to assess clinical efficacy of vaccine boosted CCP are warranted, especially since 15-20% of current donations in the US are from previously infected vaccine-boosted donors.

## INTRODUCTION

During the first waves of COVID-19 in 2020 effective therapies were lacking, so the medical community in the United States (US) and globally resorted to the use of COVID-19 convalescent plasma (CCP) from recovered patients as passive immunotherapy in hospitalized patients^1^,^2^. The widespread collection and transfusion of CCP in the US was facilitated by an Expanded Access Program approved by FDA and supported by BARDA in mid-2020, followed by FDA granting Emergency Use Authorization (EUA) status to high titer CCP in the fall of 2020. However, based on further findings from observational studies, the beneficial effects of CCP were limited to units with high titers of neutralizing antibodies (nAbs) administered early after infection, and efficacy was not confirmed by recent large randomized trials^3^. For example, the Clinical Trial of COVID-19 Convalescent Plasma in Outpatients (C3PO) study showed no significant beneficial effect in mildly symptomatic treated patients compared to controls^4^, with similar findings in the CONCOR-1 study in hospitalized patients^5^; both were halted early for futility.

Consequently, CCP use was virtually discontinued in the US during the spring of 2021, in concurrence with increased vaccination rates and availability of effective but costly monoclonal antibody (mAb) therapeutics. However, as new variants of SARS-CoV-2 emerged and spread, especially the B.1.617.2 delta variant that is resistant to neutralization by some mAbs^6^ and is the main strain involved of hospitalizations and deaths in non-vaccinated patients as well as breakthrough infections in vaccinated people at this time^7^, there is renewed interest in CCP. Although multiple studies have reported the positive impact of vaccination (including single doses or mRNA vaccines) on boosting binding and neutralizing antibodies in previously infected patients^8^, it is unclear if vaccination of previously infected CCP or regular allogeneic donors will provide CCP units with enhanced efficacy including against rapidly spreading variants of concern (VOC).

## MATERIALS AND METHODS

### Sample collection and coding

Eleven paired pre- and post-vaccination samples were provided by Dr. Thomas Gniadek, at NorthShore Medical Group, Evanston, IL. The samples had been collected between January and March 2021 from previously infected individuals CCP donors. The collection time after the first dose of the vaccine ranged from 8 to 58 days. All testing was performed on coded, anonymized samples.

### Antibody characterization

Samples were tested at Vitalant Research Institute (VRI; San Francisco, CA) to measure the levels of total Ig and IgG antibodies against the S1 domain of the SARS-CoV-2 spike antigen, in vitro binding and ACE2 inhibition against S1, N and RBD, and neutralization titers. Additional tests were conducted to assess binding and neutralization activity against different SARS-CoV2 S1 variants (B.1.1.7 - alpha, B.1.351 - beta, P.1-gamma, D614G, B.1.617-delta, B.1.427-epsilon).

Samples were first screened on the Ortho Vitros instrument (Ortho-Clinical Diagnostics, Inc., Rochester, NY) using SARS-CoV2 Ig total (COV2T) and IgG (COV2G) assays against the Spike S1 protein of the virus, as previously described^9^. Since most post-vaccination samples reached the upper limit of signal-to-cutoff (S/CO) values reported for these assays, they were further tested following dilutions (10- and 100-fold), with derivation of S/CO ratios based on reactivity levels multiplied by the dilution factors.

To assess binding and ACE2 blocking activity against ancestral SARS-CoV-2 and variants, samples were also tested on the MSD platform (MesoScale Discovery, Rockville, MD) V-PLEX SARS-CoV-2 Panel 6 Kit, (IgG binding and ACE2 inhibition) which includes SARS-CoV-2 N, SARS-CoV-2 S1 RBD, SARS-CoV-2 Spike agains the ancestral Wuhan-Hu-1, SARS-CoV-2 Spike (P.1), SARS-CoV-2 Spike (D614G), SARS-CoV-2 Spike (B.1.351), SARS-CoV-2 Spike (B.1.1.7). Briefly, plates were first incubated for 30 min with a blocking solution, then diluted samples were added, along with controls and calibrators, and incubated for 2 hours. After washing, the Detection Antibody Solution was added and incubated for 1 hour, after which the plates were washed again, followed by addition of Read Buffer, and analyzed on the MESO QuickPlex SQ 120. For direct binding serology on the MSD platform, samples were diluted 1:25,000, whereas for ACE2 inhibition dilution was 1:500. Results were reported as Arbitrary Units (AU)/ml for serology and as % inhibition compared to the negative control for the ACE2 inhibition.

In-vitro SARS-CoV-2 RVPN was performed using lentivirus-based vectors (Integral Molecular, Philadelphia, PA). Briefly, renilla luciferase RVPs bearing Spike from the ancestral strain Wuhan-Hu-1, B.1.1.7, B.1.351, P.1, B.1.1.7+E484K, B.1.617.2, and B.1.427 were first titrated on 293T/ACE2/TMPRSS2 cells. Virus concentration was then normalized and virus was incubated with equal volumes four-fold serial dilutions of heat-inactivated serum beginning at a 1 in 20 (to give a final dilution of 1 in 40). After 1 hour incubation at 37°C, 2 × 10^4^ 293T/ACE2/TMPRSS2 cells was added to each well. Plates were incubated for 3 days at 37°C and analyzed for renilla expression as per manufacturer’s instructions (Renilla-GLO, Promega, Madison, WI). Results were calculated as percent of no serum controls and dose response curves produced in Prism 9 (GraphPad) were generated to calculate 50% neutralization titers (NT_50_).

### Statistical analysis

Ortho Vitros IgG and Ig Total dilutions data were compared used matched, non-parametric Friedman test ANOVA. Pre- and post-vaccination data for each of the variants were analyzed using the Kruksal-Wallis test ANOVA, followed by Dunn’s multiple comparisons test. All tests were performed on GraphPad Prism 9.0.0.

## RESULTS

### Ortho Vitros IgG and Ig Total Binding

Antibody levels measured on the Ortho Vitros IgG assay were significantly lower before vaccination (mean S/CO 9.09, 95% CI 5.63-12.6) than after vaccination (mean S/CO 25.2, 95% CI 24.4-25.9) (Fig. 1). Similarly, the Ortho Vitros Total Ig levels were lower before the vaccination (mean S/CO 396, 95% CI 175-616) compared to the post-vaccination (mean S/CO 1197, 95% CI 1093-1301). Both assays reached the upper limit of detection for each of the assays when tested according to the EUA instructions for use, prompting additional testing following serial dilution. After dilution, the S/CO for the Total Ig post-vaccination showed an average S/CO of 12,670 (95% CI 10,917-14,423) at 1:10 and S/CO of 67,524 (95% CI 39,837-95,211) at 1:100 after adjusting for the dilution factor. for the IgG assay the post-vaccination average S/CO was 193 (95% CI 169-217) at 1:10 and average S/CO 545 (95% CI 275-816) at 1:100, after adjusting for the dilution factors.

**Fig. 1.**
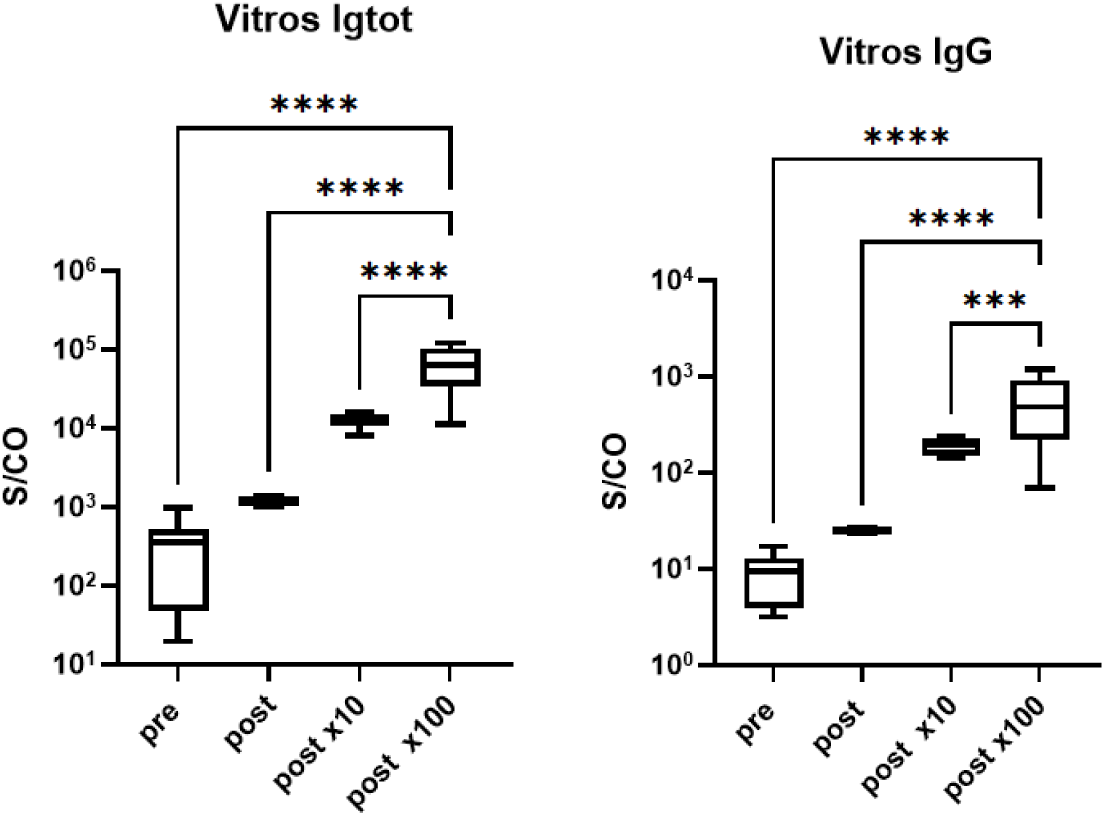
Binding antibodies levels measured on the Ortho Vitros IgG and total Ig. The total Ig S/CO (signal/cut-off) increases after vaccination, reaching the limit of detection of the assay (∼1000-1500 S/CO). After 10-fold and 100-fold dilutions, the values increase 10^2^ - 10^3^. (***=0.0005, ****= <0.0001)

### MSD IgG binding and ACE2 inhibition

The MSD serology array showed that the post-vaccination samples had an increased average AU/ml for the anti-spike and anti-RBD binding antibodies, but not for the anti-nucleocapsid, which, indeed, decreased slightly (ratio post-vaccination/pre-vaccination = 0.887) (Fig. 2). RBD binding antibodies increased the most, with a ratio of post-vaccination/pre-vaccination values of 196.3. For the SARS-CoV-2 variants, the post-vaccination levels of binding antibodies on the MSD platform increased significantly (P<0.001) across all the variants tested compared to the pre-vaccination samples, with a ratio of post-vaccination/pre-vaccination mean AU/ml between 134.1 (P1) and 78.95 (D614G). The increased levels were not significantly different among variants.

**Fig. 2.**
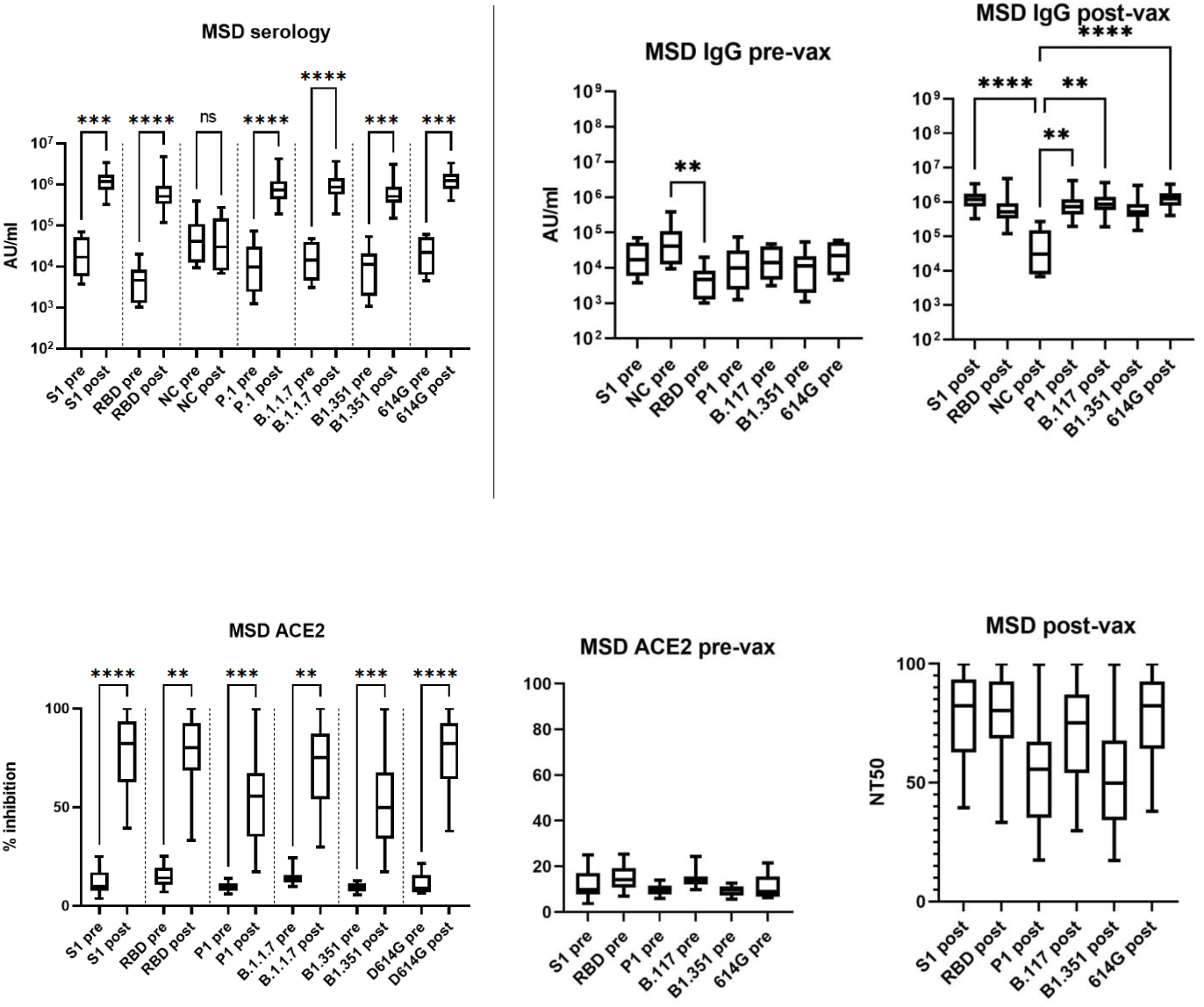
Binding IgG and ACE2 inhibition measured on the MSD V-Plex. The AU/ml IgG increases for all the post-vaccination samples, for both S1 and RBD of the Wuhan-Hu-1, but not for NC. Antibodies against all the variants increase similarly after vaccination. The percentage of ACE2 inhibition increases for all the post-vaccination samples, for Wuhan-Hu-1 S1 and RBS, and all the variants tested. There is no significance difference among all the variants, neither before, nor after vaccination. (**= 0.001, ***=0.0005, ****= <0.0001).

The ACE-2 inhibition assay showed similar trajectories following vaccination (Fig. 2). All samples showed low neutralization activity against each of the variants following natural infection, with no difference among variants before vaccination (mean=12.23%, ±2.29). After vaccination levels rose 4- to 7-fold, and similarly among all variants. The ACE2 inhibitory activity of the antibodies following vaccination of the CCP donors against all variants was on average 67.90% ± 10.74.

### Variant-specific RVPN

RVPN assays were used on pre- and post-vaccination samples to measure their ability to inhibit virus entry. All the pre-vaccination samples showed neutralizing activity due to natural infection, especially for the Wuhan-Hu-1 strain, with a mean NT50 of 547, which was significantly higher than the B1.1.7+ E484K, B.1.351 and B.617.2 (Fig. 3). After vaccination, the neutralizing activity increased significantly (p<0.001) against all the virus variants, with no significant differences in the levels of boosting between the variants. The average NT50 following vaccination was 10,831 ± 2,000. Spearman correlation analysis between the RVPN and MSD ACE2 inhibition for the VOCs, showed that the results of the two assays were strongly correlated, with r=0.8587 (95% CI 0.78-0.91).

**Fig. 3.**
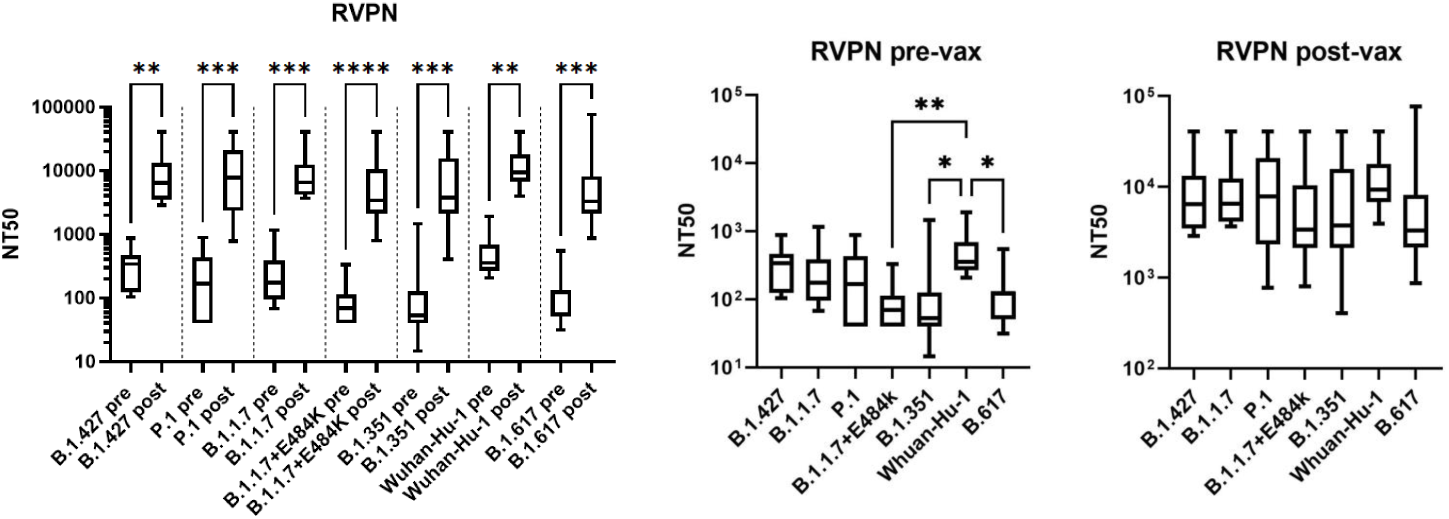
Pseudovirus neutralization measured by RVPN. The NT50 was higher against the Wuhan-Hu-1 pseudovirus before vaccination, compared to the other variants B1.1.7+ E484K, B.1.351 and B.617.2. After vaccination, the NT50 were significantly higher and comparable across variants. (*=0.05, **= 0.001, ***=0.0005).

## DISCUSSION

Despite the emergence of VOCs with decreased susceptibility to vaccine-induced immune responses ^10, 6^, vaccination still offers significant levels of protection from hospitalization and severe disease ^11^. However, following the advent of the delta (B1.617.2) VOC as the dominant circulating variant, there has been a dramatically increased surge in hospitalizations and deaths among non-vaccinated persons and reduced level of protection from infection in vaccinated individuals ^12, 11^. Several studies have reported that vaccination of previously infected individuals significantly boosts anti-SARS-CoV-2 antibody titers, and indeed one dose of the normal two-dose BNT162b2 or mRNA-1273 regimens is sufficient to boost immunity^13, 14,15^. We therefore assessed the ability of vaccination to boost anti-SARS-CoV-2 antibody levels in previously infected CCP donors, and importantly determined whether the titers of neutralizing antibody against VOCs may indicate potential value in CCP specifically collected from vaccinated, previously infected donors.

Paired pre-vaccination and post-vaccination convalescent plasma samples were tested on 5 different assays to assess antibodies levels and activity against the ancestral and variants SARS-CoV-2. As expected, both Ortho Vitros S1 IgG and total Ig S/CO levels approached or exceeded the upper limit of detection for previously infected, vaccinated subjects, making it difficult to clearly assess the real levels of antibodies. This issue is relevant in the light of the WHO international standards used for antibodies detection assays, which were originally based on naturally infected individual, while based on our dilutional studies it is clear that vaccination after natural infection synergically boosts the antibodies levels resulting in extremely high levels of so-called “hybrid immunity” ^8, 16, 17^.

Neutralizing antibodies have been proposed as correlates of protection for SARS-CoV-2^18, 19^ and the finding that nAbs elicited by previous infection and vaccination are able to neutralize different VOCs and VOIs in *vitro* is reassuring. The pre-vaccination CCP donor samples were able to neutralize the ancestral Whuhan-Hu-1 more than any other variants, but after vaccination all variants were neutralized similarly, as others have reported^16^, even against cross-clade pan-sarbecovirus^20^.

These data confirm that while natural infection provides a basic level of protection, one or two doses of vaccine are highly efficacious at boosting binding antibody reactivity and nAb titers, and also increase protection against reinfection, especially from the new variants^16,21^. It appears that vaccination following prior infection elicits higher levels of circulating neutralizing Ab, while natural infection stimulates longer B cells maturation, that eventually respond with more potent and broader Ab following a subsequent infection or vaccination^22^. Initial infections of non-vaccinated and not previously infected persons are associated with the highest risk of hospitalization and death, while vaccination of previously infected persons has proven to be safe and is associated with both protection from reinfection and increased breath of immunity and capacity to derive broadly reactive and potent monoclonal antibodies.

## Data Availability

All data produced in the present study are available upon reasonable request to the authors

## ACKNOWLEDGEMENTS

We would like to thank Paul Contestable at Ortho Clinical Diagnostic and Jim Wilbur at MesoScale Diagnostics for providing reagents for the study.

